# Metabolic correlates of late midlife cognitive function: findings from the 1946 British Birth Cohort

**DOI:** 10.1101/2020.11.23.20236463

**Authors:** Rebecca Green, Jodie Lord, Jin Xu, Jane Maddock, Min Kim, Richard Dobson, Cristina Legido-Quigley, Andrew Wong, Marcus Richards, Petroula Proitsi

**Affiliations:** Institute of Psychology, Psychiatry & Neuroscience, King’s College London, London, UK; UK National Institute for Health Research (NIHR) Maudsley Biomedical Research Centre, South London and Maudsley Hospital, London; Institute of Pharmaceutical Science, King’s College London, London, UK; MRC Unit for Lifelong Health & Ageing at UCL, University College London, London, UK; Steno Diabetes Center Copenhagen, Gentofte, Denmark

## Abstract

Investigating associations between metabolites and late midlife cognitive function could reveal potential markers and mechanistic insights relevant to early dementia. Here, we aimed to systematically explore the metabolic underpinnings of cognitive outcomes across the 7^th^ decade of life, while untangling influencing life course factors. Using levels of 1019 metabolites detected by liquid chromatography-mass spectrometry and quantified at age 60-64, we evaluated relationships between metabolites and cognitive outcomes in the British 1946 Birth Cohort (N=1740). We conducted pathway and network analyses to allow for greater insight into underlying mechanisms, and sequentially adjusted for life course factors including social factors, childhood cognition and lifestyle influences. After adjusting for multiple tests, 155 metabolites, 10 pathways and 5 network modules demonstrated relationships with cognitive outcomes. Integrating these, we identified thirty-five “hub” metabolites that were influential in their module and associated with our outcomes. Notably, we report relationships between a module comprised of acylcarnitines and processing speed that were independent of life course factors, revealing palmitoylcarnitine as a hub (final model: ß =-0.10, 95%CI =-0.15--0.052, p=5.99×10^−5^). Two modules additionally demonstrated associations with several cognitive outcomes that were partly explained by life course factors: one enriched in nucleosides and amino acids, and another in vitamin A and C metabolites. Our other findings, including a module enriched in sphingolipid pathways, were entirely explained by life course factors - particularly social factors and childhood cognition. These results highlight potential metabolic mechanisms underlying cognitive function in late midlife, suggesting marker candidates and life course relationships for further study.

## Introduction

Cognitive function in late midlife is indicative of future cognitive trajectories and risk of dementia (1). As dementia is proposed to have a long prodrome, where pathology is accumulating but clinical criteria are not yet met, there presents a promising window to prevent or delay pathology (1). However, a lack of clinically significant symptoms impedes our ability to identify individuals for potential risk-reduction and treatment strategies. As such, our understanding of early disease mechanisms are not well established and no effective disease-modifying treatments are currently used in clinic (2).

Comprehensive longitudinal studies are required to detect early mechanisms and markers preceding diagnosis, for which studying metabolic correlates may be fruitful. Metabolites, such as fatty acids and amino acids, are low molecular weight compounds derived from cellular metabolism. Lying in closest proximity to the phenotype, they represent upstream biological systems (e.g. genetics, transcriptomics, proteomics) as well as environmental and lifestyle influences, allowing for a holistic insight into the physiological status of an individual (3). Additionally, they are accessible and potentially modifiable, and thus present as promising targets of intervention (4).

The biological relevance of metabolic alterations in cognitive function and dementia has been established. Contextually, genome-wide association studies have highlighted enrichment in lipid metabolism pathways in the genetic underpinnings of Alzheimer’s disease (AD) (5). Further, many studies have linked metabolites to cognitive function and AD, consistently highlighting species such as sphingolipids, phospholipids, fatty acids, cholesterol and amino acids (6–11), although replication of specific metabolites has proved challenging.

Looking at associations of single metabolites can allow for granular insights into the molecular underpinnings of cognitive outcomes. However, metabolites are highly correlated, and biological processes are likely to involve a coordinated effort of many metabolites. As such, investigating the involvement of groups of interrelated metabolites, for example through pathways and networks, may enhance biological interpretation. One such approach is weighted gene correlation network analysis (WGCNA), whereby data are organised into modules based on pairwise correlations (12–14). Relationships between modules and outcomes can be subsequently explored, and metabolites that are highly connected in their module (“hub” metabolites) can be identified. As these metabolites are highly influential in module structure and likely to play key roles in biological function, they present as promising marker candidates for further study.

A number of studies, including ours, have applied WGCNA to identify molecular profiles associated with AD and AD endophenotypes (15–17). However, studies linking metabolites to cognitive outcomes have typically been directed towards clinical phenotypes where irreversible damage has already occurred. To our knowledge, the molecular correlates of cognitive function relevant to this prodromal window – late midlife – are yet to be explored using a network approach, and the influence of life course factors has not been previously considered. It is hoped that this could highlight independent associations as well as suggest relationships for further study – a potentially invaluable layer in untangling early pathology.

Using the Medical Research Council (MRC) National Survey of Health and Disease (NSHD) – the British 1946 Birth Cohort – we aimed to comprehensively investigate associations between metabolites and cognitive function in late midlife using a life course approach (Fig 1). Previously, levels of 233 metabolites and their association with cognitive function were explored in this cohort (6). Since that publication, we now have levels of >1000 metabolites quantified, as well as availability of the Addenbrooke’s Cognitive Examination (ACE-III), a comprehensive measure of cognitive state that is also able to screen for cognitive impairment and dementia (18). This provides a good opportunity to delineate pathways and networks associated with cognitive outcomes in late midlife. Integrating the depth and breadth of metabolite-level, pathway-level and network-level approaches, we aimed to identify functionally significant metabolites that may show merit as markers of early pathology. With lifelong information available, we explored the influence of life course factors to untangle these associations further.

**Fig 1.**
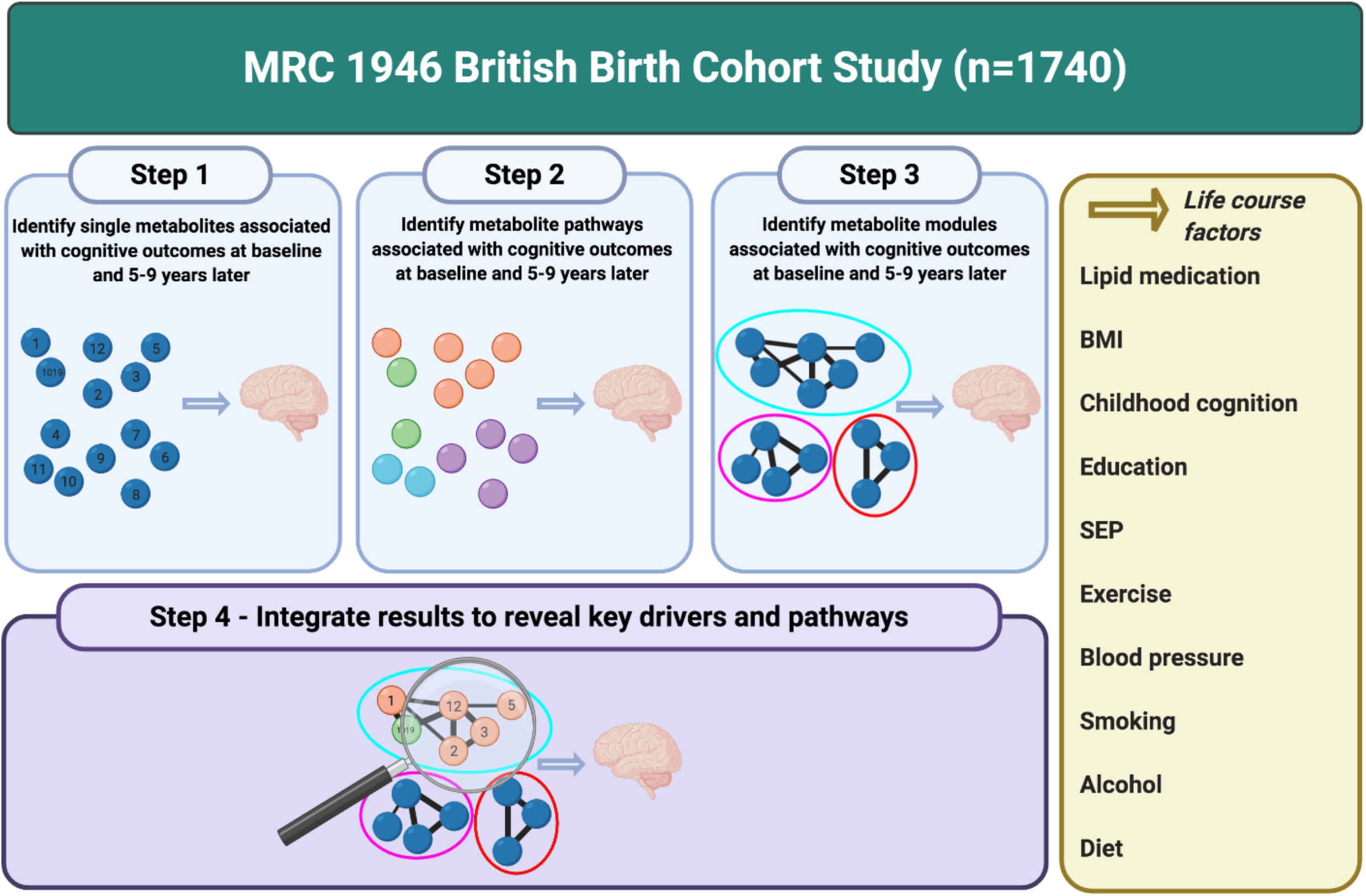
Overview of study workflow. Created with BioRender.com.

## Materials and methods

### 2.1 Participants

The MRC NSHD originally consisted of 5362 participants born in mainland Britain during one week of March in 1946 (19). Twenty-four waves of data have been collected since birth, with the most recent follow-ups at ages 60-64 (n=2229) and 68-69 (n=2149). The study sample remains broadly representative of the British-born population at the same age (19,20). Participants with full cognition, metabolite and blood clinic data at age 60-64 were included for this analysis (N=1740) (see Supplementary Fig 1).

Ethical approval was obtained from the Multicentre Research Ethics Committee (for data collections up to 2010), and the Scotland A Research Ethics Committee (14/SS/1009) and Queen Square Research Ethics Committee (13/LO/1073) (for data collections between 2014-15). Research was conducted in accordance with the Declaration of Helsinki and participants provided written informed consent at each wave.

### 2.2 Metabolomics

Blood samples were collected during the nurse visit at age 60-64 (96% fasted). Samples were aliquoted and stored at -80°C.

Levels of 1401 plasma metabolites were profiled by *Metabolon Inc* (Durham, NC, USA) using Ultrahigh Performance Liquid Chromatography-Tandem Mass Spectrometry (UPLC-MS/MS) (Supplementary Methods). Metabolites were assigned to nine families and further organised into pathways by *Metabolon* (see Supplementary Table 1).

**Table 1.** Biological family, pathway and key results for significant metabolites in the fully adjusted model (model 4)

| Metabolite | Family | Pathway | Model 1 | Model 4 | $\beta$ change from Model 1 to 4 (%) |
| --- | --- | --- | --- | --- | --- |
| X – 17676 | Unknown | Unknown | **STM (69y)**<br>$\beta$ = -0.14<br>95%CI = -0.19 to -0.089<br>$p$ = $5.29 \times 10^{-8}$<br>**DM (60-64y)**<br>$\beta$ = -0.11<br>95%CI = -0.15 to -0.061<br>$p$ = $5.45 \times 10^{-6}$ | **STM (69y)**<br>$\beta$ = -0.11<br>95%CI = -0.15 to -0.06<br>$p$ = $4.84 \times 10^{-6}$<br>**DM (60-64y)**<br>$\beta$ = -0.082<br>95%CI = -0.12 to -0.04<br>$p$ = $1.11 \times 10^{-4}$ | **STM (69y)**<br>$\beta$ change = -24.69%<br>**DM (60-64y)**<br>$\beta$ change = -23.56% |
| Palmitoylcarnitine (C16) | Lipid | Fatty Acid Metabolism (Acyl Carnitine) | **PS (60-64y)**<br>$\beta$ = -0.11<br>95%CI = -0.16 to -0.062<br>$p$ = $9.19 \times 10^{-6}$ | **PS (60-64y)**<br>$\beta$ = -0.10<br>95%CI = -0.15 to -0.052<br>$p$ = $5.99 \times 10^{-5}$ | **PS (60-64y)**<br>$\beta$ change = -9.49% |
| Margaroylcarnitine (C17)* | Lipid | Fatty Acid Metabolism (Acyl Carnitine) | **PS (60-64y)**<br>$\beta$ = -0.067<br>95%CI = -0.11 to 0.020<br>$p$ = $5.40 \times 10^{-3}$ | **PS (60-64y)**<br>$\beta$ = -0.095<br>95%CI = -0.14 to -0.048<br>$p$ = $8.33 \times 10^{-5}$ | **PS (60-64y)**<br>$\beta$ change = +40.75% |
| Imidazole propionate | Amino Acid | Histidine Metabolism | **STM (69y)**<br>$\beta$ = -0.14<br>95%CI = -0.19 to -0.091<br>$p$ = $4.98 \times 10^{-8}$ | **STM (69y)**<br>$\beta$ = -0.094<br>95%CI = -0.14 to -0.048<br>$p$ = $7.91 \times 10^{-5}$ | **STM (69y)**<br>$\beta$ change = -33.48% |

STM = short-term memory, DM = delayed memory, PS = processing speed.

Our data QC pipeline is presented in Supplementary Fig 1. Briefly, metabolites with >20% of missing data were excluded, leaving 1019 for further analysis. Remaining missing data were then imputed using k-nearest-neighbours (k=10), as recommended for LC-MS data elsewhere (21), using the *impute* package in R (22).

### 2.3 Cognitive outcomes

Cognitive outcome measures were recorded at age 60-64 and 69. Four aspects of cognitive function were assessed: short-term memory (three trial 15 item word-list; 60-64 and 69y), delayed memory (uncued delayed free recall trial; 60-64y), processing speed (letter cancellation task; 60-64 and 69y), and ACE-III (total score comprising five domains: attention and orientation, verbal fluency, memory, language and visuospatial function; 69y). Outcome measures are further detailed in Supplementary Methods.

For outcomes collected at two time points - short-term memory and processing speed - we additionally investigated change in cognition, represented by the standardised residuals of a regression model fit between scores at age 60-64 and 69.

### 2.4 Covariables

As with previous analyses (6), covariables included the following: sex, blood clinic information (age, location and fasting status), body mass index (BMI) (60-64y nurse visit), lipid medication (self-reported use in 24 hours preceding blood collection; 60-64y), childhood cognition (standardised sum of the Heim AH4 (23), Watts Vernon reading test (24) and a test of mathematical ability (24); 15y), highest level of educational attainment (no qualifications, ‘O level’, ‘A level’ or higher; 26y), childhood socioeconomic position (SEP) (Father’s current or last known occupation categorised according to the UK Registrar General’s classification; 11y), midlife SEP (own occupation categorised as specified with childhood SEP; 53y), lifetime smoking (pack years between 20 and 60-64y), alcohol intake (none, light-to-moderate, heavy; 3-5 day diet diaries at 36, 43, 53 and 60-64y), systolic blood pressure (second measurement; 60-64y), physical activity (none, 1-4 times per week, >4 times per week during month prior; 60-64y), diet (sex-specific quintiles reflecting adherence to the Dietary Approaches to Stop Hypertension (DASH) diet (25,26); diet diaries 60-64y).

*APOE* genotype was determined from blood samples collected at age 53 or 69 and analysed as described previously (27). Participants with ε2/ε4 were excluded (n=46) and *APOE* genotype was coded as homozygous ε4 (n=46), heterozygous ε4 (n=361) or non ε4 (n=1068). Genotypes were treated as continuous variables.

### 2.5 Statistical analyses

Missing covariable data were imputed using multiple imputation chained equations (*mice*) (28), resulting in fifty imputed datasets (see Supplementary Table 7 for details on missing data). All predictors and outcomes were z-standardised prior to statistical analysis to allow for direct comparisons, and all analyses were carried out in R.

#### 2.5.1 Single-metabolite analyses

Associations between metabolites and cognitive outcomes were evaluated using multiple linear regression. Regression analyses were performed on each imputed dataset, and estimates were pooled using Rubin’s rules. To investigate associations in the context of life course influences, a series of statistical models were performed:

Model 1 (basic covariables): sex, blood clinic, age at blood clinic, fasting status Model 2 (common metabolite confounders): model 1 + BMI, lipid medication Model 3 (social factors and childhood cognition): model 2 + childhood cognition, attainment, SEP (childhood and midlife) Model 4 (lifestyle influences): model 3 + blood pressure, physical activity, alcohol, smoking, diet

A Bonferroni-adjusted significance threshold was set as 0.05/ the number of principal components explaining >95% variance in metabolite data (6); p<1.15×10^−4^.

#### 2.5.2 Pathway analyses

Metabolites were assigned to pathways based on Metabolon pathway definitions. Those containing <5 metabolites were excluded, resulting in 53 pathways.

Quantitative pathway analyses were performed using an approach reported previously (29). Briefly, we derived pathway scores for each participant, representing the mean standardised expression of metabolites in the pathway. Associations between pathways and outcomes were evaluated using linear regression, adjusting for the basic (model 1) covariables listed above.

A Bonferroni-corrected significance threshold was set at 0.05/53 pathways; p<9.43×10^−4^.

#### 2.5.3 Network analyses

##### Network construction

Metabolites were first adjusted for model 1 covariables and the standardised residuals were used for subsequent analysis. We next constructed metabolite networks using WGCNA with the WGCNA package in R (12–14). Fourteen modules were identified, and module eigenvalues were computed by deriving the first principal component of the module. For further details, see Supplementary Methods.

Overrepresentation analyses using the hypergeometric test were performed on modules to identify pathways expressed more than expected by chance. For all module analyses, a Bonferroni-corrected significance threshold was set at 0.05/14 modules; p<3.57×10^−3^.

##### Regression analyses

Modules were subject to the same series of regression models listed in 2.5.1, using module eigenvalues as predictors. As modules were adjusted for model 1 covariables, these were not additionally included in model 1.

##### Module hubs

To identify hubs, we evaluated associations between metabolites and their assigned module (module membership; kME) using correlations between metabolites and module eigenvalues. Metabolites with a kME >0.65 were defined as hubs, and filtered for those identified in single-metabolite analyses.

### 2.5.4 Additional analyses

In our preliminary analysis, we investigated associations between all covariables and metabolites, and all covariables and outcomes (adjusting for model 1 covariables) (Supplementary Table 2). To further investigate whether particular covariables may be driving attenuations, we repeated single-metabolite and module regression analyses, adjusting for model 1 covariables and each additional covariable individually (Supplementary Table 3 and 4).

For significant results, analyses were rerun additionally adjusting for *APOE* genotype to investigate whether relationships were independent of *APOE* (Supplementary Table 5 and 6).

## Results

### 3.1 Participant characteristics

Complete metabolite, cognition and blood clinic data at age 60-64 were available for 1740 study participants. Repeated measures at age 69 were present for 1482 (short-term memory) and 1496 (processing speed), and ACE-III scores were present for 1255. Participant characteristics are shown in Supplementary Table 7.

### 3.2 Single-metabolite analyses

Overall, we identified 155 metabolites to be associated with a least one cognitive outcome after adjusting for multiple tests (Supplementary Fig 2). No metabolites were associated with cognitive change. Summary statistics are presented in Supplementary Table 1 and Supplementary Fig 3.

**Fig 2.**
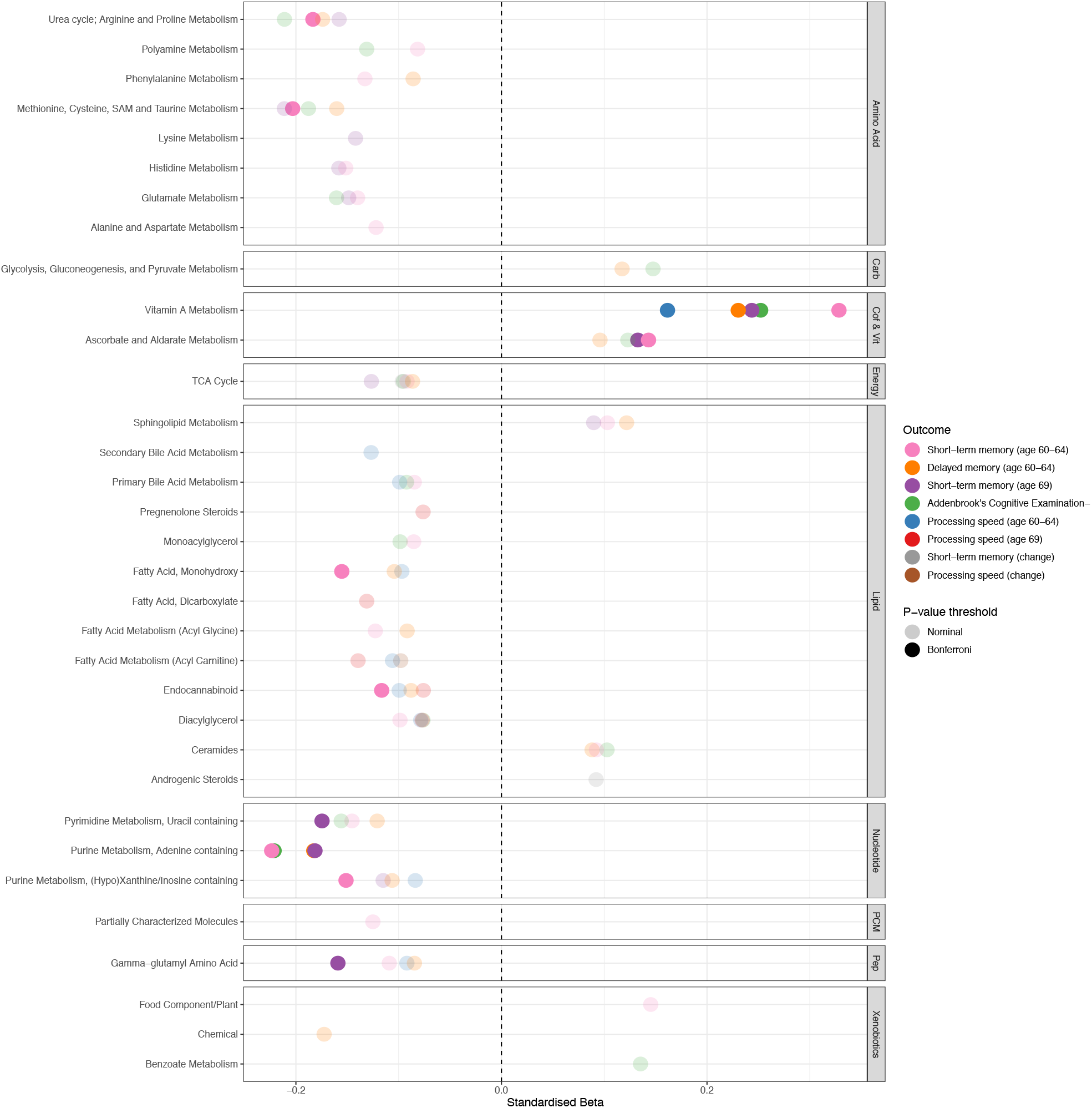
**Plot representing pathway-outcome associations, organised by metabolite family. Bonferroni-significant pathways (p<9**.**43×10**^**-4**^**) are represented by a solid fill and nominal metabolites by a faint fill (p<0**.**05). Underlying data are present in Supplementary Table 8**. *Carb = carbohydrates, Cof & Vit = cofactors and vitamins, PCM = partially characterized molecules, Pep = peptides*.

The bulk of associations attenuated after adjusting for social factors and childhood cognition (model 3), with seven metabolites remaining significant at the adjusted threshold. In the final model, four of these remained (Table 1), and 80 metabolites were nominally significant. We did not observe any significant changes following adjustment for APOE (Supplementary Table 5).

### 3.3 Pathway analyses

Results of our pathway analyses are presented in Fig 2 and Supplementary Table 8. After adjusting for multiple tests, 10 pathways showed associations with cognitive outcomes. No pathway was significant for processing speed (69y), nor for cognitive change, although some nominal associations were observed. For all other outcomes, positive relationships were seen for the Vitamin A metabolism pathway, as well as the ascorbate and aldarate metabolism pathway and short-term memory at both time points.

We observed negative relationships between the purine metabolism (adenine containing) pathway and the ACE-III and memory outcomes (short-term and delayed). Unique associations were additionally seen between various pathways belonging to amino acid, lipid, nucleotide and peptide families and short-term memory at each time point, although these pathways were nominally associated with other outcomes (Fig. 2).

### 3.4 Network analyses

WGCNA analysis identified 14 modules comprising 22-192 metabolites; five of these were associated with cognitive outcomes in model 1 at the adjusted threshold (p<3.57×10^−3^) (Fig 3A). All but one module were enriched in a biological pathway (Fig 3B and Supplementary Table 9), and no relationships were seen for cognitive change measures (p<3.57×10^−3^). Key results are presented in Fig 3 and summary statistics in Supplementary Table 10.

**Fig 3.**
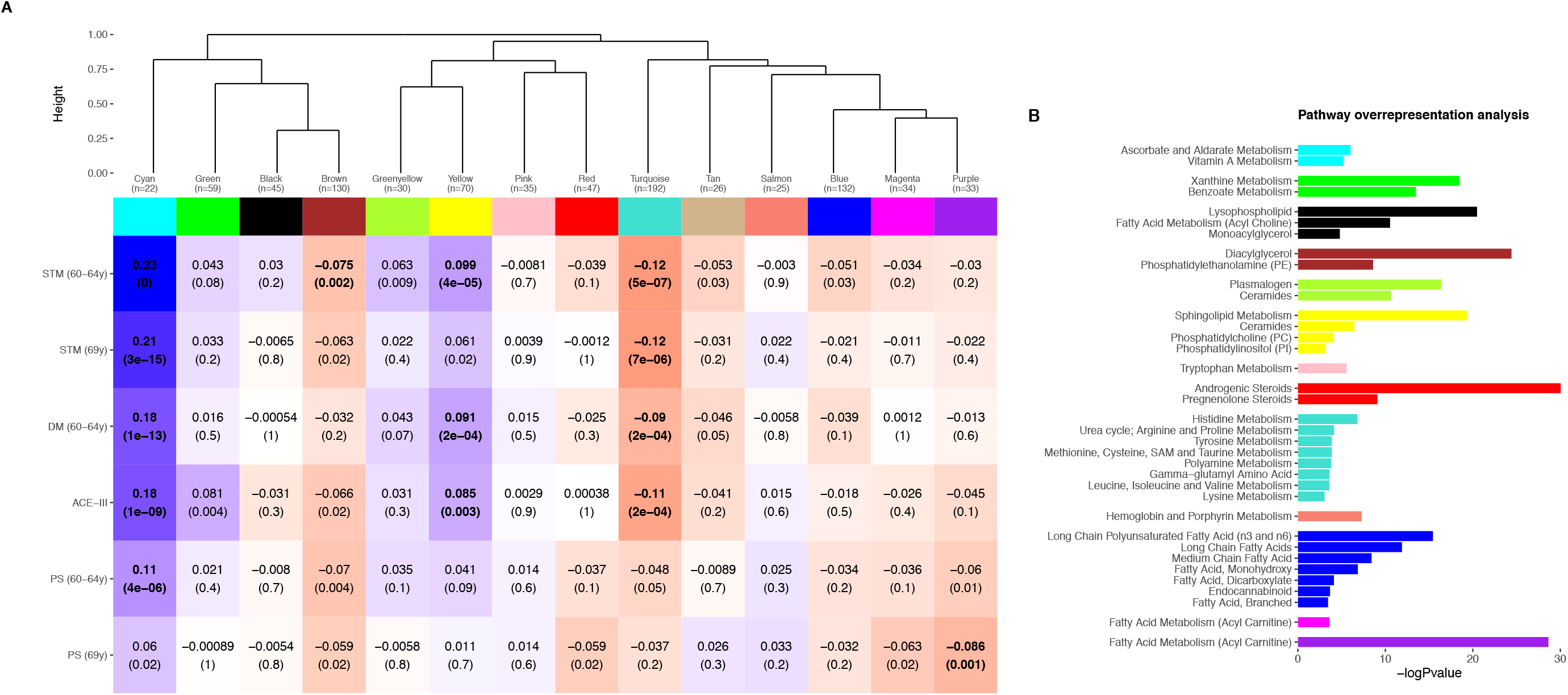
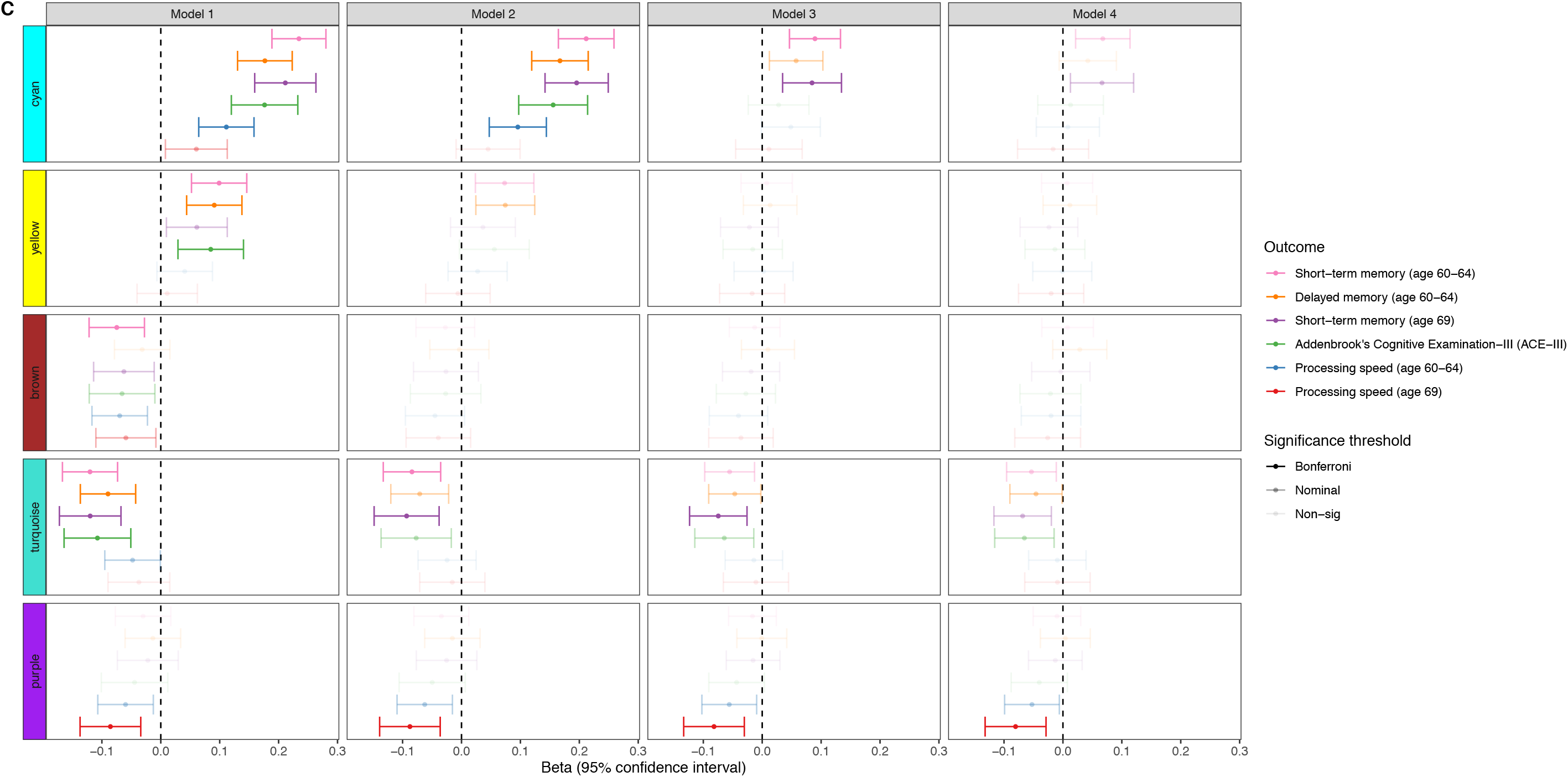
**A) Module dendrogram and heatmap of module-outcome associations. Effect sizes and unadjusted p-values are presented in the tiles and significant associations (p<3**.**57×10**^**-3**^**) are highlighted in bold. For clarity purposes, only outcomes demonstrating a Bonferroni-significant result are shown. B) Overrepresented pathways for each module. Only pathways significant at the adjusted threshold (p<9**.**43×10**^**-4**^**) are shown. P-values are unadjusted. C) Forest plot showing associations between modules and outcomes in models 1-4. Bonferroni-significant modules (p<3**.**57×10**^**-3**^**) are represented by a solid fill, nominal modules (p<0**.**05) by a fainter fill and modules that are not significant at either threshold are represented by the faintest fill. For clarity purposes, only outcomes demonstrating a Bonferroni-significant result are shown. Underlying data are present in Supplementary Table 9 and 10**. *STM = short-term memory, DM = delayed memory, ACE-III = Addenbrooke’s Cognitive Examination-III, PS = processing speed*

**Fig 4.**
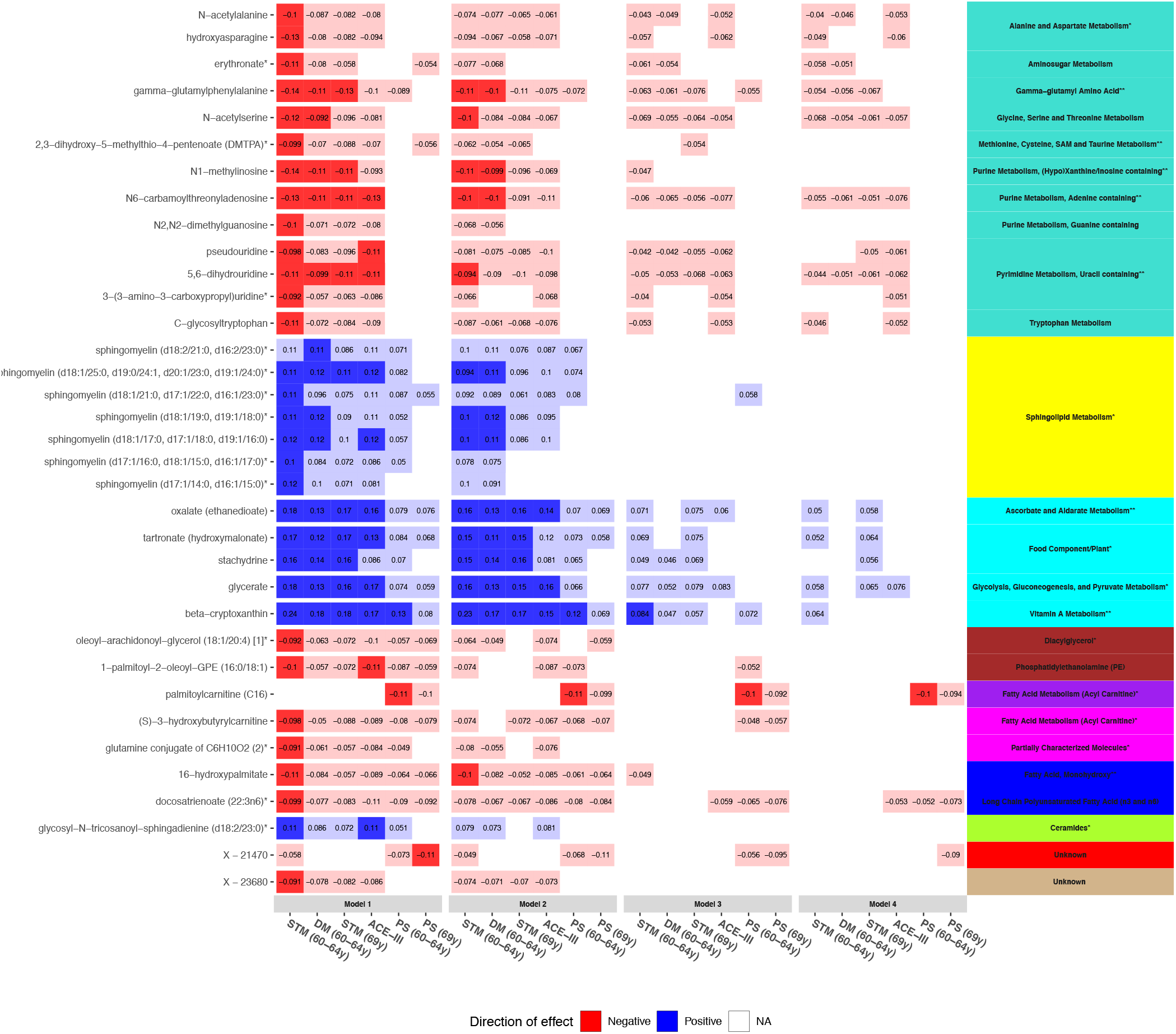
**Heatmap showing trends of associations between hub metabolites and cognitive outcomes in models 1-4. Panels on the right indicate the pathways (text) and modules (colour) represented by metabolites. Pathways are suffixed with an asterisk if they were previously identified in our pathway analyses (** = p<9**.**43×10**^**-4**^, **\* = p<0**.**05). Bonferroni-significant metabolites (p<1**.**15×10**^**-4**^**) are represented by a solid fill, nominal metabolites by a faint fill (p<0**.**05), and non-significant metabolites by no fill (p>0**.**05). Tiles are coloured by effect direction and effect sizes are noted in the centre. For clarity purposes, only outcomes demonstrating a Bonferroni-significant result are shown**.**Underlying data are present in Supplementary Table 1**. *STM = short-term memory, PS = processing speed, DM = delayed memory, ACE-III = Addenbrooke’s Cognitive Examination-III*.

#### 3.4.1 Positive relationships (cyan and yellow)

After adjusting for multiple tests, the cyan module – enriched in ascorbate and aldarate metabolism, vitamin A metabolism and food/plant consumption – was associated with all memory outcomes, processing speed (64y) and ACE-III (model 1: ß range=0.11-0.23, p range=<2.16×10^−16^-3.52×10^−6^). In the final model, no relationships were significant at the adjusted threshold, but associations for short-term memory remained nominally associated (60-64y: ß=0.068, 95%CI =0.022-0.11, p=4.10×10^−3^; 69y: ß=0.066, 95%CI =0.013-0.12, p=0.015). Overall attenuations ranged from 68.6-92.6%. Relationships were most sensitive to model 3 adjustments; our additional analysis indicated that all model 3 covariables were able to reduce associations, with childhood cognition and education resulting in the biggest attenuations (Supplementary Table 4).

We similarly found the yellow module – enriched in sphingolipid metabolism and related pathways – to display positive associations with ACE-III and age 60-64 memory outcomes at the adjusted threshold (model 1: ß range=0.085-0.10, p range=3.64×10^−5^-2.85×10^−3^). These findings were substantially or fully attenuated in the final model (87.0-116%). Relationships were sensitive to model 2 and 3 adjustments, and substantial reductions (66.3-84.8%) were seen in model 3, with childhood cognition and education resulting in the biggest attenuations (Supplementary Table 4).

#### 3.4.2 Negative relationships (turquoise, purple and brown)

The turquoise module - enriched in several amino acid metabolism pathways - displayed negative associations with ACE-III, short-term memory and delayed memory (model 1: ß range=-0.12--0.09, p range=5.23×10^−7^-2.05×10^−4^). All relationships remained at the nominal threshold in the final model, but none passed multiple testing correction: standardised betas ranged from -0.068--0.046; and p values from 0.0060-0.043. Overall attenuations ranged from 39.2-55.5%, with the majority of attenuations coming from model 2 and 3 adjustments.

Negative associations were also identified between the purple module - enriched in fatty acid (acyl carnitine) metabolism – and processing speed (age 69) (model 1: ß=-0.086, 95%CI =-0.14--0.034, p=1.15×10^−3^). This relationship remained significant and relatively stable, demonstrating an effect size reduction of 6% overall and a final effect size of ß=-0.080 (95%CI=-0.13--0.029, p=2.33×10^−3^). Similar effect directions were detected for both processing speed (60-64y) and change in processing speed at the nominal threshold (60-64y (model 4): ß=-0.053, p=0.027; change (model 4): ß=-0.065, p=0.016).

Finally, we identified the brown module - enriched in diacylglycerol and phosphatidylethanolamine pathways - to be negatively associated with short-term memory at age 60-64 (model 1: ß =-0.075, 95%CI =-0.12--0.028, p=1.85×10^−3^). Associations attenuated in model 2, which appeared to be mainly driven by BMI (Supplementary Table 4). Subsequent model adjustments resulted in full attenuation.

Results were largely unchanged after adjusting for APOE (Supplementary Table 6).

#### 3.4.3 Hub metabolites

Thirty-five metabolites identified in single-metabolite analyses were revealed to be hubs, and 28 of these belonged to the five modules that were associated with cognitive outcomes (Fig 6 and Supplementary Table 1). Hubs further represented eight of the 10 pathways identified in pathway analyses: gamma-glutamyl amino acid; methionine, cysteine, SAM and taurine metabolism; purine metabolism, (hypo)xanthine/inosine containing; purine metabolism, adenine containing; pyrimidine metabolism, uracil containing; ascorbate and aldarate metabolism; vitamin A metabolism; and fatty acid (monohydroxy). In model 4, one hub belonging to the purple module – palmitoylcarnitine (C16) – remained significant at the adjusted threshold and 17 were nominally significant.

## 4. Discussion

Using the British 1946 Birth Cohort, we systematically evaluated the metabolic correlates of cognitive function in late midlife while untangling influencing life course factors. We identified 155 metabolites, 10 pathways and five network modules to show associations with cognitive outcomes. Integrating these, 35 hub metabolites were revealed to show potential as markers for further study. Some relationships were independent of life course influences, highlighting unique metabolic mechanisms underlying cognitive outcomes. However, consistent with our previous analyses in the MRC 1946 (6), as well as a previous lipidomics study in the Lothian Birth Cohort (30), many were sensitive to childhood cognition and education, suggesting important considerations for future studies.

Most notably, we report independent relationships between the purple module - enriched in medium and long chain acylcarnitines - and processing speed, with our pathway analyses largely in support of this. These relationships were specific to processing speed, indicating a possible mechanism unique to this outcome. One metabolite, C16 (a long chain acylcarnitine), appeared to be a key driver of these associations, suggesting a potential candidate for further investigation.

Biologically, medium and long chain acylcarnitines are derivatives of fatty acid metabolism and known to be pivotal in mitochondrial fatty acid oxidation (31). Increased abundances in serum have thus been regarded as proxies for mitochondrial dysfunction and impairments in subsequent energy production (31). More specifically, C16 has also been linked to the induction and regulation of apoptotic events (32). Both apoptosis and mitochondrial dysfunction have been implicated in neurodegeneration, indicating a plausible biological mechanism behind our observations (33,34).

Perturbations in acylcarnitine levels have been reported in early cognitive impairments and AD (7,35,36), as well as other outcomes, such as insulin resistance (37,38), obesity (38) and cardiovascular disease (39). Although these traits show influence on cognitive function, our associations remained following adjustment for related factors such as blood pressure, BMI and lipid medication. Taken together, this presents the possibility that alterations may reflect metabolic dysfunction, conferring vulnerability to adverse health outcomes, including decline in processing speed. Future studies will seek to establish whether these changes lie on the causal pathway.

We also discovered a module comprised of nucleotides and amino acids, turquoise, to demonstrate negative associations with the ACE-III and memory outcomes. As the largest module, it was enriched in several pathways, including three which were brought to our attention in pathway analyses (histidine metabolism; methionine, cysteine, SAM and taurine metabolism; gamma-glutamyl amino acid metabolism). Purine and pyrimidine metabolism pathways were additionally represented by module hubs and further corroborated by pathway analyses. Relationships were partly explained by life course factors, but remained significant at the nominal threshold in the final model.

Module hubs were amino acids and nucleosides that were unified by the presence of modifications, and included in these were several markers of RNA turnover and oxidative stress (40–42). Interestingly, the top driver of the module was 2,3-dihydroxy-5-methylthio-4-pentenoic acid (DMTPA), belonging to the methionine, cysteine, SAM and taurine metabolism pathway. This pathway has a key role in post-transcriptional and post-translational modifications, including the sourcing of methyl and aminocarboxypropyl groups demonstrated in some module hubs (43,44), suggesting possible relationships linking these metabolites.

With key roles in the expression, function and stability of molecules, RNA and amino acid modifications are able to regulate a multitude of biological and pathological processes (45,46). Interestingly, many of these module hubs have been consistently reported together in adverse outcomes, including hypertension (47), chronic kidney disease (48,49), inflammation (50), mortality (51), and worse processing speed (10). Here, we extend this to late midlife cognitive outcomes, as well as a clinical measure of cognitive state - the ACE-III.

The widespread associations of these metabolites together in adverse outcomes presents the possibility that they could reflect converging molecular aetiologies. A previous study hypothesised they may represent an ‘accelerated ageing’ phenotype and suggested modified nucleosides to be a marker of tissue breakdown and oxidative stress (52), which have been implicated in worse cognitive outcomes (53). Lending some support for this, factors previously related to oxidative stress and accelerated ageing, such as BMI and smoking, were associated with the module, but none were able to suitably explain relationships. It will be interesting to dissect this in future studies.

We reported relationships between sphingolipids and improved cognitive function which were entirely explained by life course factors. In our basic analysis, the yellow module – enriched in several pathways related to sphingolipid metabolism – showed associations with age 60-64 memory outcomes and ACE-III, with pathway analysis largely corroborating this. Module relationships were attenuated by model 3, and particularly sensitive to childhood cognition and education. Given the attention sphingolipid metabolism has received in the neurodegeneration field, this warrants further consideration.

Sphingolipids are a lipid family comprised of sphingomyelins, ceramides and glycosphingolipids, and are present in large quantities in the CNS (54). Forming important components of cell membranes, they are highly dynamic and are thought to display crucial roles in cognitive development and function (55). Previous research has implicated disturbances in sphingolipid balance in cognitive development (55,56), function (55), ageing (55,57) and AD (54), but replication of specific sphingolipids has proved challenging. This could be due to fluctuations as a function of age or pathology, or inconsistencies in incorporating life course factors in analytical models could be a contributor.

Given observational findings linking sphingolipids and cognitive function at several stages of the life course, there exists several possibilities behind the sensitivity of relationships to adjustment for model 3 covariables, and particularly childhood cognition and education. For childhood cognition, this suggests confounding through reverse causation. In the case of education, this may plausibly confound associations, resulting in spurious associations for cognitive outcomes. However, without longitudinal metabolite data, we cannot rule out the possibility that later life sphingolipid levels may be a proxy for those of earlier life. In this regard, childhood cognition and education could mediate associations between early life sphingolipid levels and later life cognitive function; or perhaps they may be linked by common genetic or environmental causes. Unravelling the precise nature of these relationships will be crucial in understanding the importance of sphingolipids in cognitive outcomes.

Finally, a module enriched in vitamin A and C metabolites, cyan, showed positive associations with most cognitive outcomes. Our model 1 results suggested a ubiquitous positive role of vitamin A metabolites, displaying the largest overall effects across all stages of analysis. Nevertheless, associations were sensitive to adjustment for life course factors, namely social factors and childhood cognition. In the final model, relationships were largely explained for processing speed and ACE-III, but remained at the nominal threshold for short-term memory at both time points.

Oxidative stress is thought to be involved in the pathogenesis of neurodegenerative diseases and is characterised by an overabundance of reactive oxygen species, initiating a host of deleterious effects (58). Antioxidants, including vitamin A and C metabolites, may inhibit such processes through scavenging of these species (59). Due to this, their involvement in ageing, cognitive decline and AD has been discussed, and all five module hubs have shown protective effects in small studies investigating cognitive impairments (60–62). Nevertheless, contributions are debated, with epidemiological studies showing conflicting results (63,64).

In the final model, relationships were largely explained for processing speed and ACE-III, but nominal residual relationships remained for short-term memory at both time points, with age 60-64 just shy of the adjusted threshold. Although these relationships showed marked attenuations, this indicates that there may be some independent effects specific to short-term memory at these ages. As discussed with previously, attenuations suggest confounding by social factors and childhood cognition, or may reflect relationships we are not able to capture without longitudinal metabolite data. Further research is required to understand these relationships in greater depth.

Findings should be considered in light of several limitations. First, our results may be subject to residual confounding and a lack of longitudinal metabolomic data precludes the investigation into lifelong relationships and directionality; our next step will be to interrogate causality through methods such as Mendelian Randomisation. Next, cognitive change measures were curated from data collected within a narrow time window, which could explain the lack of relationships observed. Change measures were also represented by residualised change scores, which can be subject to bias, and our findings should be interpreted with this in caution. Finally, as seen with many cohort studies, individuals remaining in the study at this stage were generally of higher cognitive ability in childhood and more socially advantaged compared to the sample initially recruited at birth. Further, the study sample were ethnically homogenous. For generalisation, it will be paramount to replicate this work in more diverse populations.

In summary, we conducted one of the largest LC-MS studies to date on cognitive outcomes in late midlife and are the first to evaluate systems-level associations in the context of life course factors. We integrated metabolites, pathways and networks, offering biological interpretation while retaining granularity, and highlighted molecular mechanisms underlying cognitive outcomes in late midlife. Our results illustrate the importance of incorporating life course influences, with many relationships largely explained by childhood cognition and education. Finally, we identified several metabolites (e.g. C16) that were both pivotal to module function and associated with our outcomes, presenting as potential marker candidates for additional study.

## Data Availability

Data are stored and held by the MRC Unit of Lifelong Health & Ageing, UCL. Requests for data can be made by contacting the unit directly.

## Acknowledgements

PP is funded by Alzheimer’s Research UK and RG by the National Institute for Health Research (NIHR) Biomedical Research Centre at South London and Maudsley NHS Foundation Trust and King’s College London.

This paper represents independent research part-funded by the National Institute for Health Research (NIHR) Biomedical Research Centre at South London and Maudsley NHS Foundation Trust and King’s College London. The views expressed are those of the author(s) and not necessarily those of the NHS, the NIHR or the Department of Health and Social Care. This work was further supported by the UK Medical Research Council which provides core funding for the MRC National Survey of Health and Development (MC_UU_12019/06).

We thank NSHD study members for their lifelong participation and past and present members of the NSHD study team who helped to collect the data. We also thank ACE Mobile for providing a customised version of the ACE-III for NSHD.

## Supplementary information is available at the MP’s website

