## Supplementary Legends for "Metabolic correlates of late midlife cognitive function: findings from the 1946 British Birth Cohort"

**Supplementary Table 1. Full single-metabolite analyses results (models 1-4), including module membership and hub status.**

**Supplementary Table 2. Linear regression analyses results for the association of each covariable against each predictor and outcome. All analyses are adjusted for sex, age at blood collection and blood collection centre.**

**Supplementary Table 3. Linear regression analyses results for the associations between metabolites and outcomes, after adjusting for each covariable individually. All analyses are adjusted for sex, age at blood collection and blood collection centre.**

**Supplementary Table 4. Linear regression analyses results for the associations between modules and outcomes, after adjusting for each covariable individually.**

**Supplementary Table 5. Linear regression analyses results for the associations between metabolites and outcomes with and without adjusting for APOE. The maximum N with APOE genotype was used.**

**Supplementary Table 6. Linear regression analyses results for the associations between metabolites and outcomes with and without adjusting for APOE. The maximum N with APOE genotype was used.**

**Supplementary Table 7. Clinical and demographic characteristics of participants of the MRC 1946 British Birth Cohort study.**

**Supplementary Table 8. Pathway analyses results.**

**Supplementary Table 9. Module overrepresentation analysis results.**

**Supplementary Table 10. Full module analyses results (models 1-4).**

**Supplementary Fig 1. Flow chart depicting metabolomic data quality control.**

**Supplementary Fig 2. Upset plot depicting the number of metabolites associated with each outcome and combination of outcomes at the adjusted threshold, split by metabolite family. Outcomes are shown in the matrix, with shaded circles demonstrating those represented by the bar chart above. Where more than one outcome is indicated, lines further highlight these intersections. Bars are coloured by metabolite family and total metabolite counts are displayed on top of each bar, alongside the corresponding percentage proportion of all significant metabolites identified across all outcomes. As no metabolites were significant at the adjusted threshold for change outcomes, these were not included. (Left) Barplot showing the number of metabolites identified overall for each outcome. Bars are coloured by metabolite family and counts are displayed on the top of each bar. This plot was produced using (66). Underlying data are present in Supplementary Table 1.** *ACE-III = Addenbrooke's Cognitive Examination-III.*

**Supplementary Fig 3. Heat map showing trends of associations between the 155 metabolites and cognitive outcomes in models 1-4, organised by metabolite family. Bonferroni-significant metabolites ($p<1.15x10^{-4}$) are represented by a solid fill, nominal metabolites by a faint fill ($p<0.05$), and non-significant metabolites by no fill ($p>0.05$).** *Carb = carbohydrates, Cof & Vit = cofactors & vitamins, En = energy, PCM = partially characterised molecules, Pep = peptides, ACE-III = Addenbrooke's Cognitive Examination-III, DM = delayed memory, STM = short-term memory, PS = processing speed*

**Supplementary Fig 4. Scale free topology and mean connectivity plots for weighted gene coexpression network analysis.**

**Supplementary Methods. Supplementary methods**
