## Supplementary Methods for "Metabolic correlates of late midlife cognitive function: findings from the 1946 British Birth Cohort"

### Metabolomic profiling

**(Information provided by Metabolon Inc)**

**Sample Accessioning:** Following receipt, samples were inventoried and immediately stored at -80°C. Each sample received was accessioned into the Metabolon LIMS system and was assigned by the LIMS a unique identifier that was associated with the original source identifier only. This identifier was used to track all sample handling, tasks, results, etc. The samples (and all derived aliquots) were tracked by the LIMS system. All portions of any sample were automatically assigned their own unique identifiers by the LIMS when a new task was created; the relationship of these samples was also tracked. All samples were maintained at -80°C until processed.

**Sample Preparation:** Samples were prepared using the automated MicroLab STAR® system from Hamilton Company. Several recovery standards were added prior to the first step in the extraction process for QC purposes. To remove protein, dissociate small molecules bound to protein or trapped in the precipitated protein matrix, and to recover chemically diverse metabolites, proteins were precipitated with methanol under vigorous shaking for 2 min (Glen Mills GenoGrinder 2000) followed by centrifugation. The resulting extract was divided into five fractions: two for analysis by two separate reverse phase (RP)/UPLC-MS/MS methods with positive ion mode electrospray ionization (ESI), one for analysis by RP/UPLC-MS/MS with negative ion mode ESI, one for analysis by HILIC/UPLC-MS/MS with negative ion mode ESI, and one sample was reserved for backup. Samples were placed briefly on a TurboVap®

(Zymark) to remove the organic solvent. The sample extracts were stored overnight under nitrogen before preparation for analysis.

**QA/QC:** Several types of controls were analyzed in concert with the experimental samples: a pooled matrix sample generated by taking a small volume of each experimental sample (or alternatively, use of a pool of well-characterized human plasma) served as a technical replicate throughout the data set; extracted water samples served as process blanks; and a cocktail of QC standards that were carefully chosen not to interfere with the measurement of endogenous compounds were spiked into every analyzed sample, allowed instrument performance monitoring and aided chromatographic alignment.Instrument variability was determined by calculating the median relative standard deviation (RSD) for the standards that were added to each sample prior to injection into the mass spectrometers. Overall process variability was determined by calculating the median RSD for all endogenous metabolites (i.e., non-instrument standards) present in 100% of the pooled matrix samples. Experimental samples were randomized across the platform run with QC samples spaced evenly among the injections.

**Ultrahigh Performance Liquid Chromatography-Tandem Mass Spectroscopy (UPLC-MS/MS):** All methods utilized a Waters ACQUITY ultra-performance liquid chromatography (UPLC) and a Thermo Scientific Q-Exactive high resolution/accurate mass spectrometer interfaced with a heated electrospray ionization (HESI-II) source and Orbitrap mass analyzer operated at 35,000 mass resolution. The sample extract was dried then reconstituted in solvents compatible to each of the four methods. Each reconstitution solvent contained a series of standards at fixed concentrations to ensure injection and chromatographic consistency. One aliquot was analyzed using acidic positive ion conditions, chromatographically optimized for more hydrophilic compounds. In this method, the extract was gradient eluted from a C18 column (Waters UPLC BEH C18-2.1x100 mm, 1.7 µm) using water and methanol, containing 0.05% perfluoropentanoic acid (PFPA) and 0.1% formic acid (FA). Another aliquot was also analyzed using acidic positive ion conditions, however it was chromatographically optimized for more hydrophobic compounds. In this method, the extract was gradient eluted from the same afore mentioned C18 column using methanol, acetonitrile, water, 0.05% PFPA and 0.01% FA and was operated at an overall higher organic content. Another aliquot was analyzed using basic negative ion optimized conditions using a separate dedicated C18 column. The basic extracts were gradient eluted from the column using methanol and water, however with 6.5mM Ammonium Bicarbonate at pH 8. The fourth aliquot was analyzed via negative ionization following elution from a HILIC column (Waters UPLC BEH Amide 2.1x150 mm, 1.7 µm) using a gradient consisting of water and acetonitrile with 10mM Ammonium Formate, pH 10.8. The MS analysis alternated between MS and data-dependent $MS^n$ scans using dynamic exclusion. The scan range varied slighted between methods but covered 70-1000 m/z. Raw data files are archived and extracted as described below.

**Data Extraction and Compound Identification:** Raw data was extracted, peak-identified and QC processed using Metabolon's hardware and software. These systems are built on a web-service platform utilizing Microsoft's .NET technologies, which run on high-performance application servers and fiber-channel storage arrays in clusters to provide active failover and load-balancing. Compounds were identified by comparison to library entries of purified standards or recurrent unknown entities. Metabolon maintains a library based on authenticated standards that contains the retention time/index (RI), mass to charge ratio (*m/z)*, and chromatographic data (including MS/MS spectral data) on all molecules present in the library. Furthermore, biochemical identifications are based on three criteria: retention index within a narrow RI window of the proposed identification, accurate mass match to the library +/- 10 ppm, and the MS/MS forward and reverse scores between the experimental data and authentic standards. The MS/MS scores are based on a comparison of the ions present in the experimental spectrum to the ions present in the library spectrum. While there may be similarities between these molecules based on one of these factors, the use of all three data points can be utilized to distinguish and differentiate biochemicals. More than 3300 commercially available purified standard compounds have been acquired and registered into LIMS for analysis on all platforms for determination of their analytical characteristics. Additional mass spectral entries have been created for structurally unnamed biochemicals, which have been identified by virtue of their recurrent nature (both chromatographic and mass spectral). These compounds have the potential to be identified by future acquisition of a matching purified standard or by classical structural analysis.

**Curation:** A variety of curation procedures were carried out to ensure that a high quality data set was made available for statistical analysis and data interpretation. The QC and curation processes were designed to ensure accurate and consistent identification of true chemical entities, and to remove those representing system artifacts, mis-assignments, and background noise. Metabolon data analysts use proprietary visualization and interpretation software to confirm the consistency of peak identification among the various samples. Library matches for each compound were checked for each sample and corrected if necessary.

**Metabolite Quantification and Data Normalization:** Peaks were quantified using area-under-the-curve. A data normalization step was performed to correct variation resulting from instrument inter-day tuning differences. Essentially, each compound was corrected in run-day blocks by registering the medians to equal one (1.00) and normalizing each data point proportionately.

#### Cognitive outcomes

Cognitive outcome measures were recorded at two ages, 60-64 and 69. Four aspects of cognitive function were assessed:

*Short-term memory (age 60-64 & 69)*

Participants were asked to recall a 15-item word list, developed by the NSHD, after being presented with each word for two seconds. The task was repeated over three trials and the number of accurately recalled words was recorded (max score=45) (1).

*Processing speed (age 60-64 & 69)*

Participants were asked to cross out the letters P and W, randomly distributed on a page containing other letters. One minute was given to complete the task and participants were scored by the number and accuracy of the letters crossed out (max score=600) (1).

*Delayed memory (age 60-64)*

After the processing speed task, an uncued delayed free recall trial was administered (1).

*Addenbrooke's Cognitive Examination-III (ACE-III) (age 69)*

The ACE-III captures cognitive state, and is also a screening tool for cognitive impairment, comprised of five domains: attention and orientation, verbal fluency, memory, language and visuospatial function. Scores represent the total over all domains (max score=100), with lower scores indicating poorer cognitive function (2).

#### Weighted gene correlation network analysis (WGCNA)

To define metabolic networks, we applied weighted gene coexpression network analysis (WGCNA) to metabolite data, using the WGCNA package in R (3–5). WGCNA is a network analysis approach that organises data into densely connected modules based on pairwise correlations, whereby data in the same module will show high connectivity and those in differing modules, low. Subsequently, the first principal component of the module (module eigenvalue) can be derived and relationships between modules and outcomes can be explored. To infer biological function, overrepresentation analyses are commonly conducted to identify enriched pathways within the module. Additionally, the function of metabolites showing the greatest modular connectivity can be interrogated (3).

Metabolites were first adjusted for model 1 covariables and the standardised residuals were used for subsequent analysis. Next, the standardised connectivity (Z.k) for each sample was computed to identify outliers, resulting in the exclusion of ten individuals with a Z.k of < -4. We then derived a pairwise correlation matrix using biweight midcorrelations between all metabolites. From this, a weighted, signed adjacency matrix was constructed by raising correlations to a soft thresholding power of 9, chosen to meet a scale-free topology threshold of >= 0.85 while maximising mean connectivity (Supplementary Fig 4). Subsequently, the adjacency matrix was transformed into a topological overlap matrix (TOM), representing the network connectivity of metabolites. Metabolites were then hierarchically clustered into a dendrogram using an average linkage method based on their dissimilarity (1-TOM), and the dendrogram was cut using a dynamic hybrid tree cutting algorithm (6) (parameters - minModuleSize=20, deepSplit=4 and mergeHeight=0.3), resulting in 15 metabolite modules. Of these, the 'grey' module, comprised of metabolites that were not assigned to any particular module, was dropped from further analysis. Module eigenvalues were computed for the remaining 14 modules.
