## Supplementary Figures for "Metabolic correlates of late midlife cognitive function: findings from the 1946 British Birth Cohort"

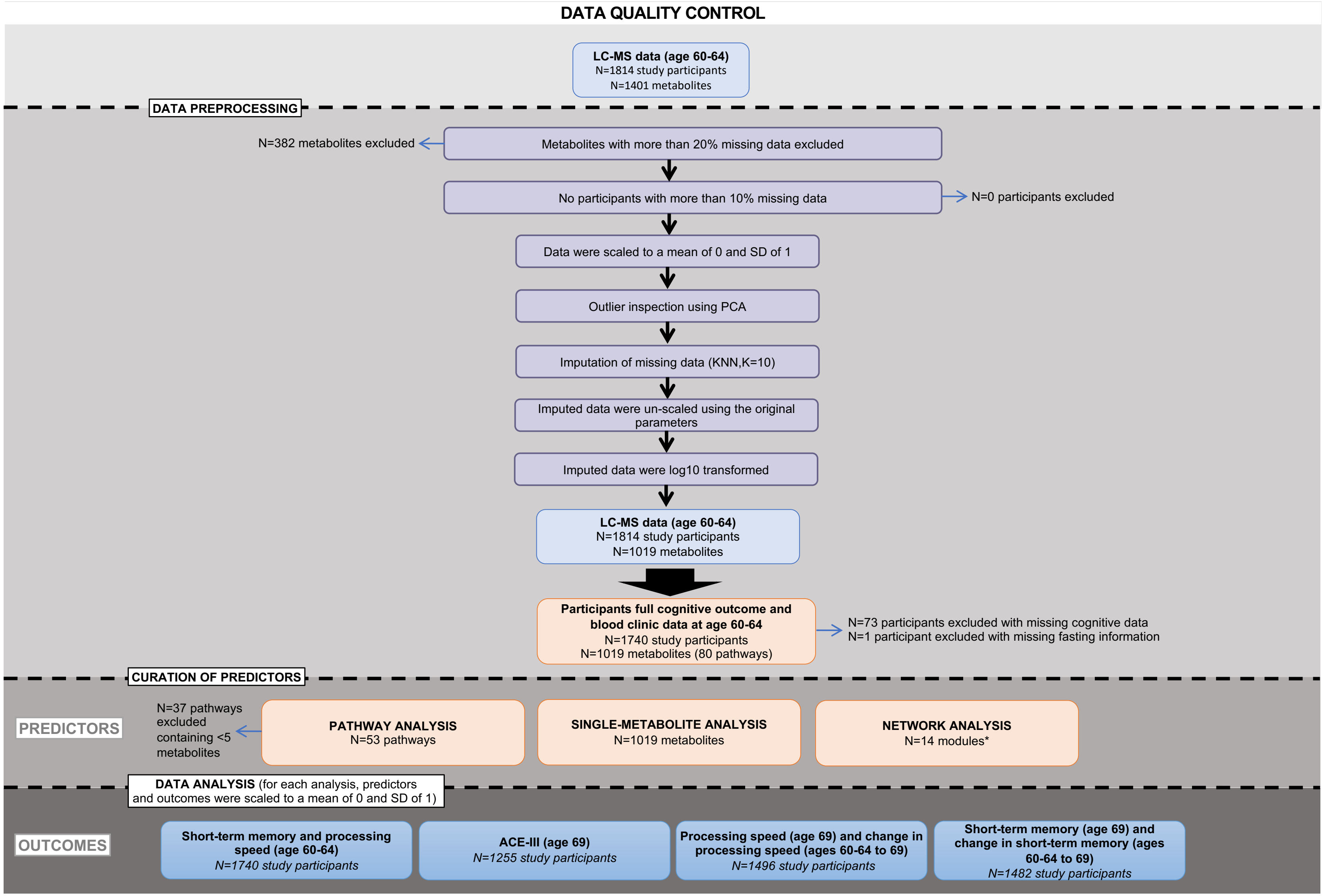


*In the data analysis step, all predictors were tested against all outcomes. *Before module curation, 10 participants with a standardised connectivity of < -4 were excluded.*

**Supplementary Fig 1.**

### Supplementary Figures

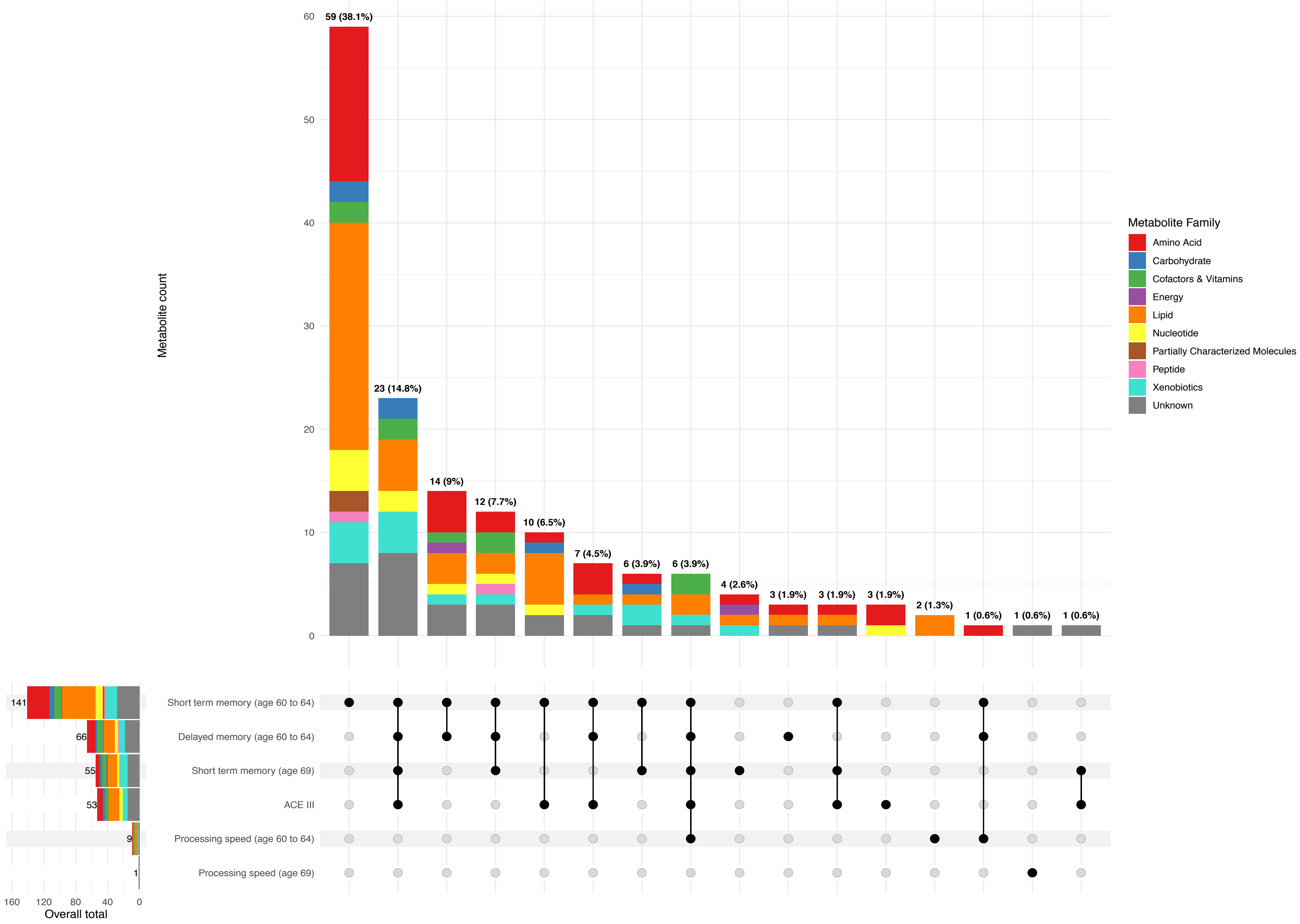


**Supplementary Fig 2.**

### Supplementary Figures

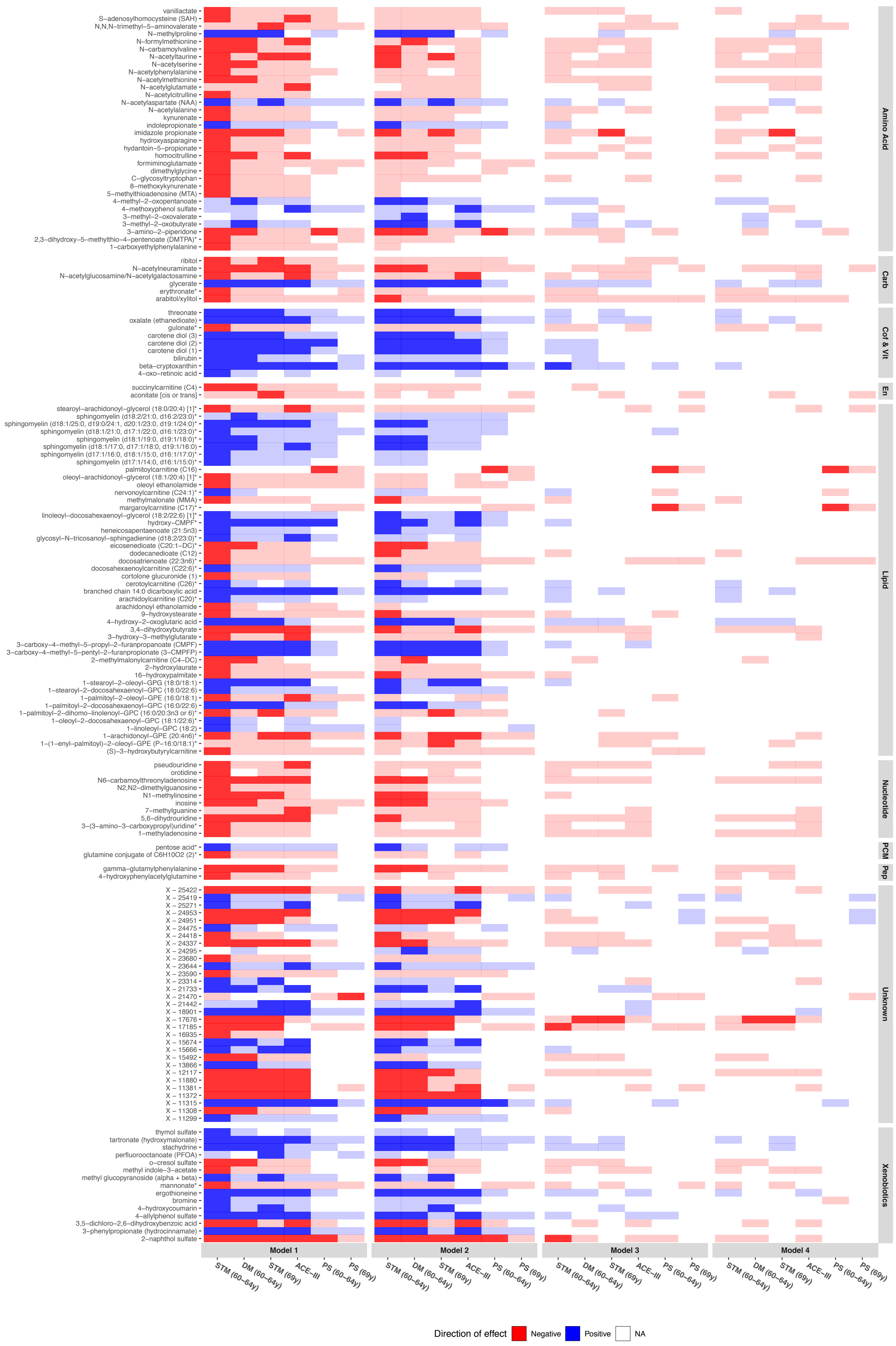


### Supplementary Fig 3.

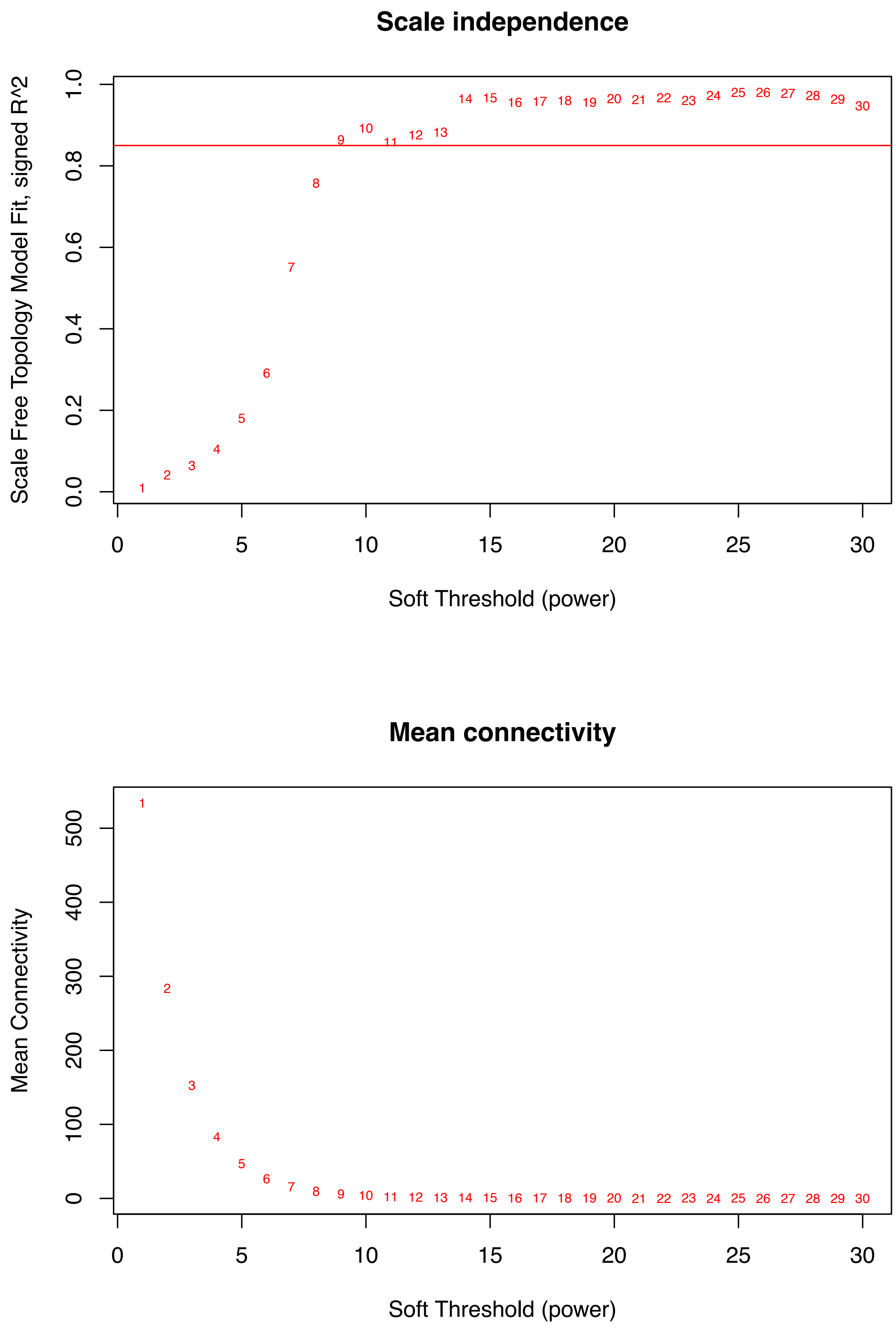


**Supplementary Fig 4.**
